# Clinical performance of a chemiluminescence SARS-CoV-2 antibody assay in a cohort of healthcare workers, blood donors and COVID-19 patients

**DOI:** 10.1101/2023.03.09.23287052

**Authors:** Giselle Rangel, Daysa Lopez, Athneris Chavarría, Laiss Mudarra, Gabrielle Britton, Alcibiades Villarreal

**Affiliations:** Institute for Scientific Research and High Technology Services (INDICASAT-AIP), City of Knowledge, Panama; School of Medical Technology, Faculty of Medicine, University of Panama (UP), Panama City, Panama; Dr. Arnulfo Arias Madrid Metropolitan Hospital Complex, Social Security Fund, Panama City, Panama; National Research System (SNI) by Panama’s National Secretariat of Science, Technology and Innovation (SENACYT), Panama City, Panama

**Keywords:** antibody test, chemiluminicense, SARS-CoV-2, COVID-19, serology

## Abstract

**Introduction:** Serological detection of antibodies against SARS-CoV-2 has become an essential tool to test vaccine efficacy and epidemiological surveillance of COVID-19. There have been limited published studies documenting the performance of SARS-CoV-2 antibody assays within hispanic populations.

**Materials and methods:** We evaluated the diagnostic performance of a chemiluminescence enzyme immunoassay (CLIA) on a set of 1,035 samples including pre-pandemic samples, healthcare workers (HCW), blood donors (BD) and COVID-19 positive confirmed by RT-PCR collected from April to December 2020.

**Results:** Through a ROC curve the CLIA test had a high diagnostic performance, with an AUC of 0.9854 (CI_95%_ 95.68-100), P <0.0001. The analysis yielded a cut-off point 0.1950, sensitivity of 98.4% (CI_95%_ 95 91.54-99.9), and specificity of 93.8% (CI_95%_ 79.8 - 98.9). The diagnostic performance was also evaluated comparing the results with those obtained using other diagnostic techniques. Substantial agreement with the lateral flow chromatography and RT-PCR tests was found, and a high level of agreement with ELISA, with %PPA of 91.3 (CI_95%_ 84.0-95.5), % NPA of 97.7 (CI_95%_ 96.3-98.6), % OPA of 97.7 (CI_95%_ 96.3-98.6) and Cohen’s kappa value of 90.4 (CI_95%_ 85.8-94.9). A logistic regression was used to determine which of the independent variables predicted reactivity to CLIA test. A higher age was associated with an odds ratio (OR) of 1.043 (CI_95%_ 1.022-1.065), while the presence of at least one chronic disease was associated with an OR of 5.649 (CI_95%_ 3.089-10.329) greater likelihood of reactivity.

**Conclusions:** CLIA test exhibited excellent performance making it a suitable test for seroprevalence surveillance at the community level.

## INTRODUCTION

Antibody detection methods as markers of prior infection with severe acute respiratory syndrome coronavirus 2 (SARS-CoV-2) have a significant role to play in the response to the COVID-19 pandemic. Chemiluminescence immunoassay (CLIA), enzyme-linked immunosorbent assay (ELISA) and lateral flow immunoassay (LFIA) are among the popular serological assays used to detect specific antibodies against the virus (1). ELISA is by far the most commonly used method (2), however, some CLIAs have several advantages over ELISA in terms of turnaround time (3), sensitivity and specificity, particularly <7 days after the onset of the disease (1),. Several commercial assays are available, but few manufacture-independent evaluations and few comparisons between assays with data from Black or Hispanic populations have been performed (4–6). Also, depending on the chosen antigen and assay protocol, SARS-CoV-2 IgG antibody test specificity may be significantly reduced in certain populations, possibly due to interference of immune responses to endemic pathogens such as other viruses or parasites (5,7,8). Over 40 novel SARS-CoV-2-specific antibody testing kits have been developed but there is lack of information regarding their relative performance with respect to the RT-PCR, the gold standard test for SARS-CoV-2 detection. The comparison between assays is hampered by the absence of accepted serological antibody gold standards, and therefore studies evaluating the concordance between assays are needed. The primary purpose of the data collected in this analysis was to estimate the performance of a rapid lateral flow chromatography test and seroprevalence of past SARS-CoV-2 infection among healthcare workers (HCW) and blood donors (BD) in the Panamanian population (9). The data presented here are a secondary analysis with the aim to evaluate the diagnostic performance of a commercial CLIA anti-SARS-CoV-2 immunoassay in serum specimens of 3 study groups and pre-pandemic samples. The sensitivity and specificity of this CLIA test was compared with RT-PCR and with other available results from serological techniques including LFIA and ELISA assays.

## MATERIALS AND METHODS

### Subjects/Materials

This is a retrospective cross-sectional clinical study of the seroprevalence of circulating antibodies to SARS-CoV-2 in hospital HCW and BD conducted between April to December 2020. The location of participants recruitment and study inclusion/exclution criteria was described previously (9). The participants were asked to complete a short questionnaire about demographic and clinical data regarding the presence of respiratory symptoms, preexisting comorbidities and suspected contact with COVID-19 positive individuals. Total anti-SARS-CoV-2 antibodies from serum specimens from 343 HCW, 258 BD, 110 COVID-19 patients who were positive to SARS-CoV-2 by RT-PCR, and 32 pre-pandemic samples (negative controls) were measured by commercial electrochemiluminescence immunoassay. Some HCW had a second blood sample (visit 2) taken at an interval of 15±5 days from the first sampling (visit 1).

The clinical study was registered with the Panama Ministry of Health (No. 1,462) and the protocol was approved by the National Research Bioethics Committee (CNBI; No. EC-CNBI-2020-03-43). All participants provided informed consent. Anonymity and confidentiality of the study participants were maintained.

### Methods

Blood samples and serum specimens were obtained and preserved using standardized protocols following good laboratory practices. Prior to analysis, all serum samples were heat-inactivated at 56°C for one hour (10).

The VITROS Immunodiagnostic Products Anti-SARS-CoV-2 Total test develops the amplified chemiluminescence method for the qualitative detection of total antibodies (IgG, IgA and IgM) against the S1 subunit of the SARS-CoV-2 S protein using the Vitros 5600 automatic system. The test was performed according to manufacturer protocol. The chemiluminescent reaction resulting from the assay is measured as a unit of light (reactivity index). Assay results greater than or equal to the cut-off point (≥1) are labeled “reactive” indicating the presence of anti-SARS-CoV-2 antibodies, and results below the cut-off point (<1) are labeled as “non-reactive” indicating the absence of anti-SARS-CoV-2 antibodies in the sample analyzed. The analytical sensitivity and specificity reported by the manufacturer, Ortho Clinical Diagnostics, is 100%, in a 95% confidence interval. Information about the manufacturer, assay commercial name, volume required, test waiting time and temperature are summarized in Supplementary Table 1.

### Statistical analysis

Data analysis was carried out to evaluate the diagnostic performance of CLIA test. The results obtained were compared against other available serological immunoassays (ELISA, LFIA tests) and RT-PCR results. The results were classified as true positives, false positives, true negatives and false negatives. The percentage of positive and negative concordance was estimated are reported with 95% confidence interval. Bivariate analyses were performed using Spearman correlation tests. Two-way ANOVA, Kaplan-Meier curve analysis and forward logistic regression analyses were used to test whether various factors were associated with reactivity, and odds ratios (OR) are reported with a 95% confidence interval. Diagnostic performance of CLIA test was evaluated with a receiver operating characteristic (ROC) curve analysis. All statistical analyses were conducted using a level of significance of p < 0.05 using SPSS (version 24) and GraphPad Prism version 8.0.0 for Windows, GraphPad Software, San Diego, California USA, www.graphpad.com

## RESULTS

### Sample distribution

The study sample consisted of 743 individuals and a total of 1,035 serum samples analyzed with CLIA platform, which included 635 samples from healthcare workers (HCW), 258 samples from blood donors (BD), 110 samples from hospitalized COVID-19 positive patients, and 32 serum samples collected in a pre-pandemic period as a COVID-19 negative control group (Figure 1).

**Figure 1.**
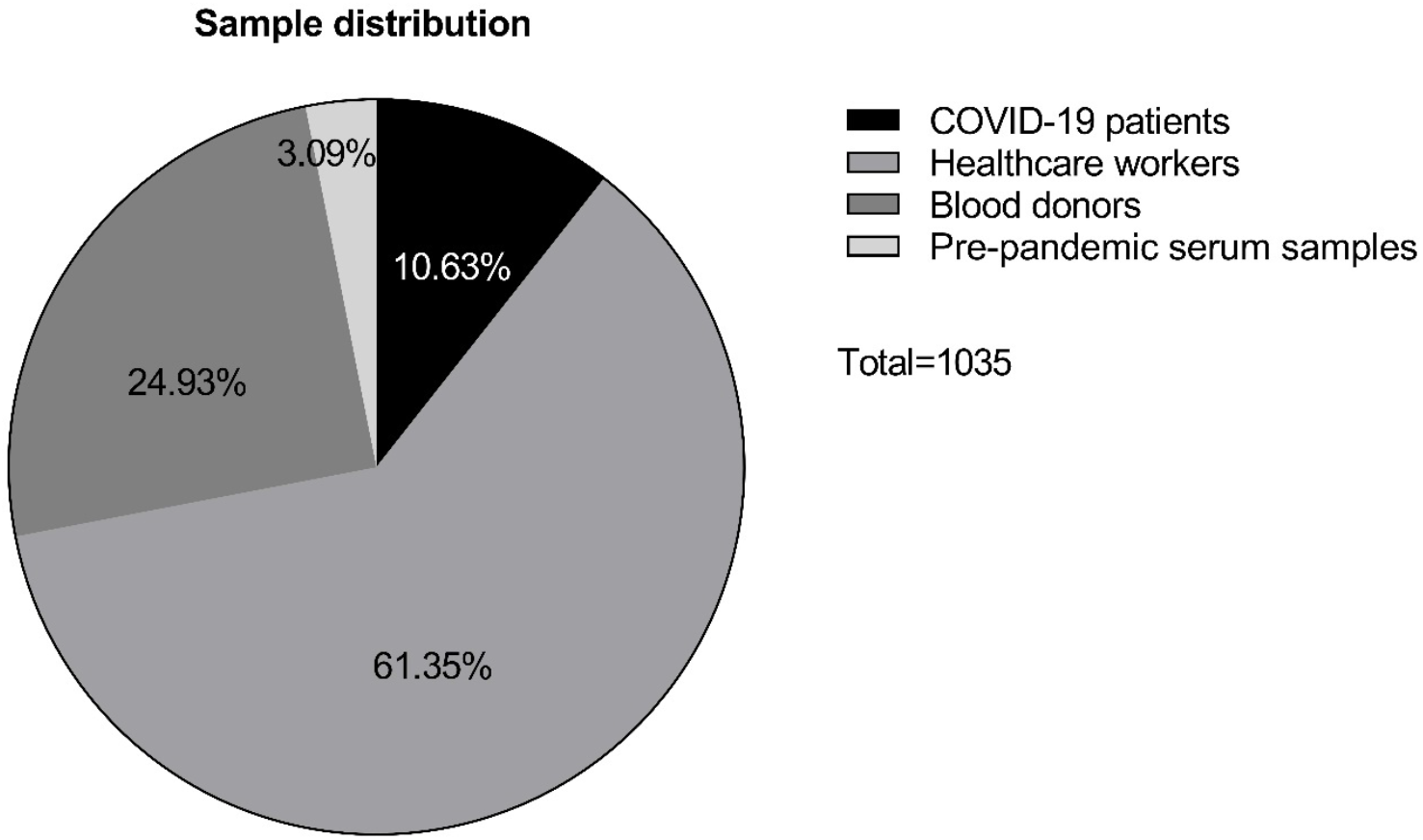
Distribution of the analyzed sample. The total sample included 635 samples from health workers (61%), 258 blood donors (25%), 110 COVID-19 positive patients (11%) and 32 pre-pandemic serum samples (3%). Numbers in the table correspond to the percentage of the total sample.

### Characterization of clinical study sample

In the total study population, sex proportion was similar among participants (supplementary table 2). Most the individuals (94.2%) in the study were within an age range of 18-64 years. Half of the participants were in an age range of 18 to 39 years (50.1%), while 5.8% were 65 years or older. Within the HCW group the proportion of women was greater (67.9%), while in BD group the proportion of men was greater (69.4%). In the group of hospitalized COVID-19 participants, the highest proportion was in the age range of 40 to 64 years (57.3%), in addition, the percentage of people aged 65 years and over was 28.2%.

The majority of the study population reported not suffering from chronic diseases (69.6%), while in hospitalized COVID-19 participants (n=110) 75.5% reported having at least one chronic disease. The most prevalent chronic diseases were hypertension (50.0%), followed by diabetes mellitus (34.6%) and renal insufficiency (13.6%). Most of the study population reported not knowing or having had contact with COVID-19 positive people (56.2%), while 72.6% of HCW participants reported being exposed to COVID-19 positive people.

### Percentage of reactivity to CLIA test

Of the total pre-pandemic samples evaluated, none showed reactivity to the test, as expected (SARS-CoV-2 negative control samples). On the other hand, of the total study sample, a percentage of reactivity to the test of 10.2% (CI_95%_ 8.5-12.2) was found. In HCW, 5 positive results were found (Table 1), representing a percentage of reactivity of 1.5% (CI_95%_ 0.5-3.4). In BD group, 9 people reactive to the test were detected, reaching a percentage of reactivity of 3.5% (CI_95%_ 1.8-6.6). In COVID-19 positive group, 80.0% (CI_95%_ 71.5-86.5) demonstrated reactivity against CLIA test. Correlation between anti-SARS-CoV-2 reactivity index and days post-diagnosis

**Table 1.** Percentage of reactivity of CLIA test to detect total anti-SARS-CoV-2 antibodies.

| Percentage<br>of reactivity | Total | HCW<br>Visit 1 | HCW<br>Visit 2 | BD | COVID-19<br>positive | Pre-pandemic<br>samples |
| --- | --- | --- | --- | --- | --- | --- |
|  | n= 1035 | n= 343 | n= 292 | n= 258 | n= 110 | n= 32 |
| Non-reactor | 929<br>(89.76%) | 338<br>(98.54%) | 288<br>(98.63%) | 249<br>(96.51%) | 22<br>(20.00%) | 32<br>(100%) |
| Reactor | 106<br>(10.24%) | 5<br>(1.46%) | 4<br>(1.37%) | 9<br>(3.49%) | 88<br>(80.00%) | 0<br>(0%) |

Comments: HCW= healthcare workers, BD= blood donors

CLIA consistently detected antibodies within the first 15 days after RT-PCR diagnosis, with 80% sensitivity. In samples collected 15 days or more after RT-PCR diagnosis, CLIA achieved 100% sensitivity. In order to determine if the CLIA test reactivity index correlated with days after RT-PCR diagnosis, we analyzed the data from COVID-19 positive participants (n= 63) and performed a Spearman correlation coefficient (r) analysis (Figure 2), where we obtained an r of 0.2895 (CI_95%_ 0.037 to 0.507), P = 0.0214.

**Figure 2.**
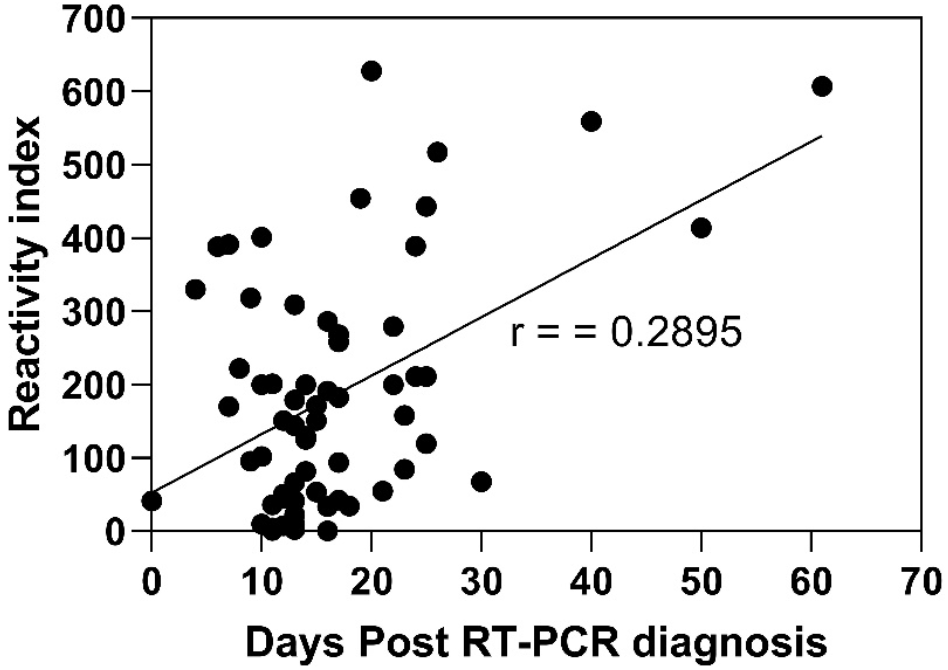
Correlation between the reactivity rate of COVID-19 positive patients according to the time elapsed since diagnosis by RT-PCR. Diagonal line represent r value.

### Association between SARS-CoV-2 seropositivity and potential risk factors

We stratified the results of CLIA test according to clinical responsibilities and risk of exposure in the work environment of HCW in three categories: A) doctors/nurses (direct exposure, high risk), B) medical/laboratory technologists (indirect exposure, medium risk) and C) administrative (not exposed, low risk), the percentages of participants by category are described in Figure 3. Also, the result was stratified according to the report of having or not having contact with COVID-19 positive people. Table 2 shows the results obtained through a two-way ANOVA and a Kaplan-Meier curve analysis. We found no significant differences between HCW individuals with respect to clinical responsibilities, nor contact with COVID-19 positive people, in relation to the results obtained with CLIA test.

**Table 2.**
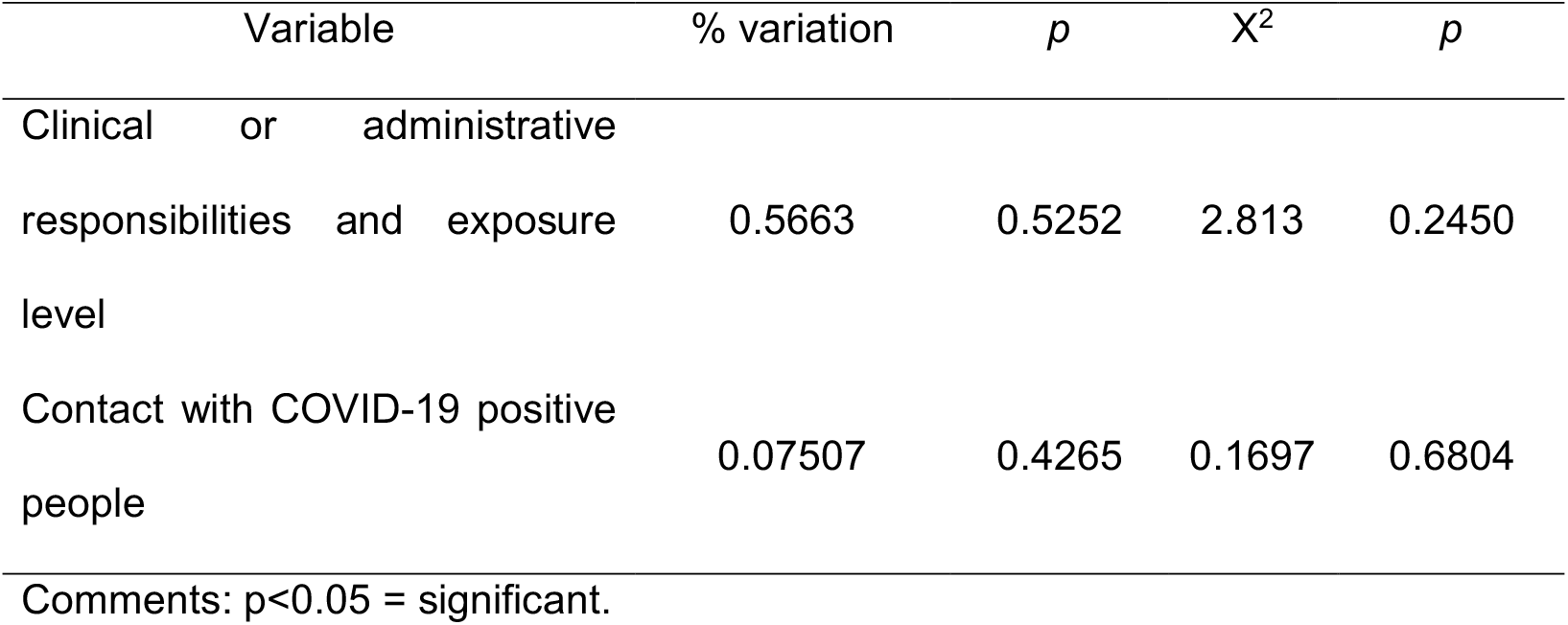
Association of risk level and contact with COVID-19 positive people, with seropositivity to the test in HCW

**Figure 3.**
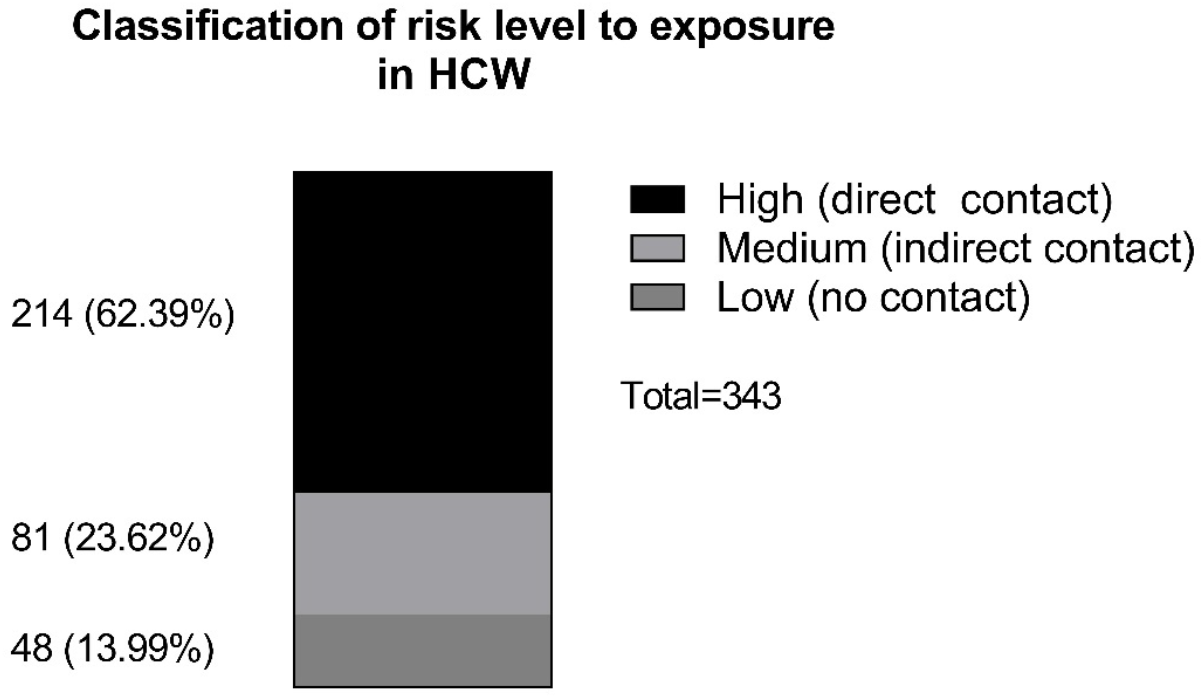
Stratification of the group of Health workers according to clinical or administrative responsibilities and level of exposure to COVID-19 patients. HCW classified as High level were those with clinical responsibilities and direct contact with suspicious patients such as doctor, nurse, nursing technician, emergency medical technician, physiotherapy technician, respiratory therapy technician, admission officer, and insurance officer. Medium level corresponded to indirect contact with COVID-19 patients such as personnel in the following areas coordination, medical technology, laboratory, assistant technician, laboratory assistant, radiology, psychology. Low level corresponded to administrative personnel such as receptionist, secretary, courier, clerk, manual worker, director, escort, general assistant, advisor, engineer, health educator, statesman, and storekeeper.

Forward logistic regression was conducted to determine which variables are predictors of reactivity. Regression results indicated that the overall model was statistically reliable in distinguishing between reactive and non-reactive test results; χ^2^(4) = 90.392, p<0.001. The model correctly classified 89.2% of the cases. Older age was associated with 1.043 (CI_95%_ 1.022-1.065) and chronic illness with 5.649 (CI_95%_ 3.089-10.329) greater likelihood of reactivity (Table 3).

**Table 3.**
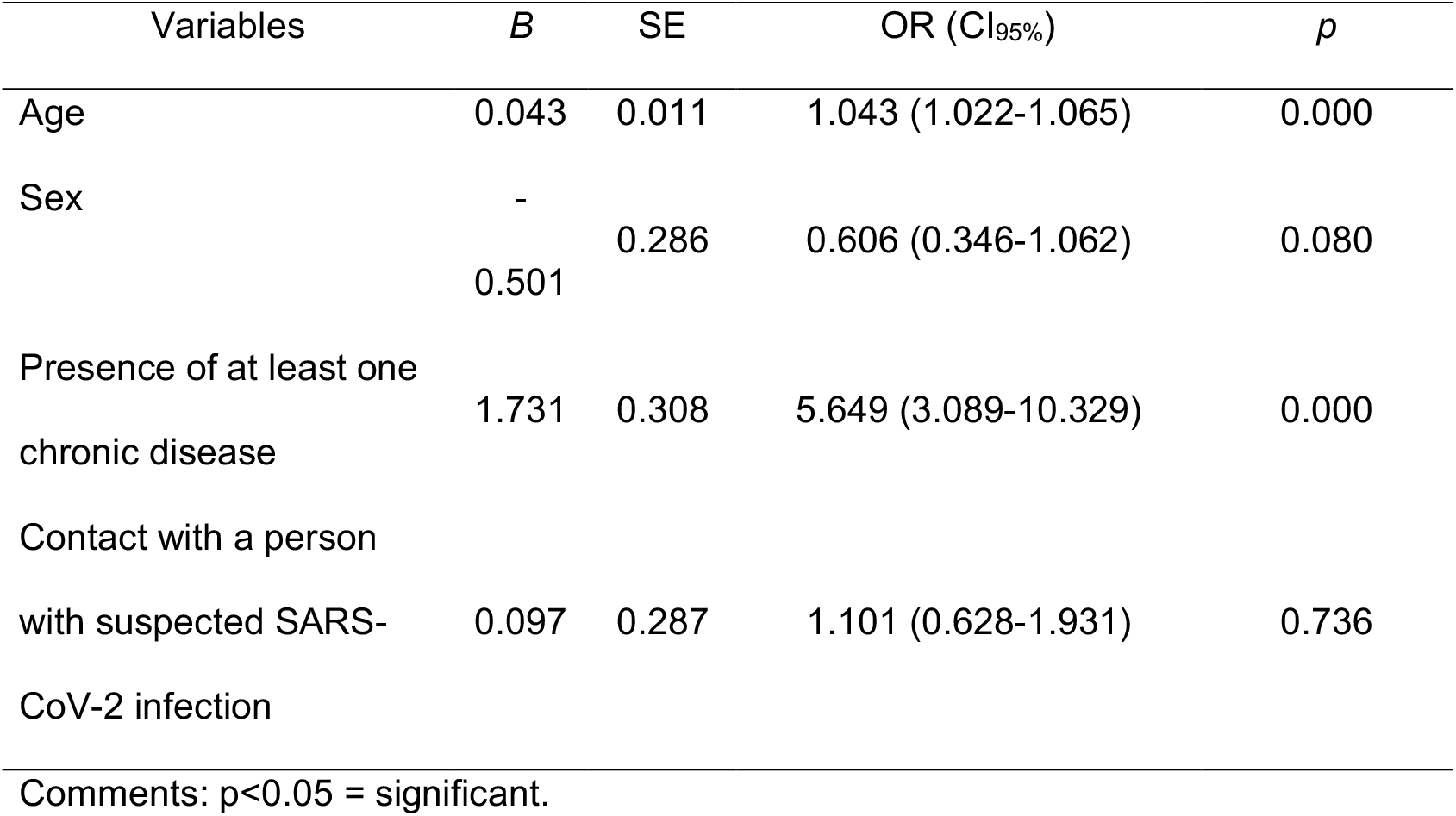
Binary logistic regression analysis of factors associated with serological reactivity to the SARS-CoV-2 virus.

### Diagnostic performance of VITROS Immunodiagnostic Products Anti-SARS-CoV-2 test

We conducted a ROC curve analysis (receiver operating characteristic) using the results obtained with the pre-pandemic samples as a negative control and those from the group of COVID-19 patients as positive samples. The results obtained are shown in Figure 4, the CLIA test demonstrated a high diagnostic performance, with an area under the curve (AUC) of 0.9854 (CI_95%_ 95.68-100), P = <0.0001. The analysis showed an optimal cut-off point > 0.1950, with a sensitivity of 98.4% (CI_95%_ 91.5-99.9), specificity of 93.8% (CI_95%_ 79.8-98.9).

**Figure 4.**
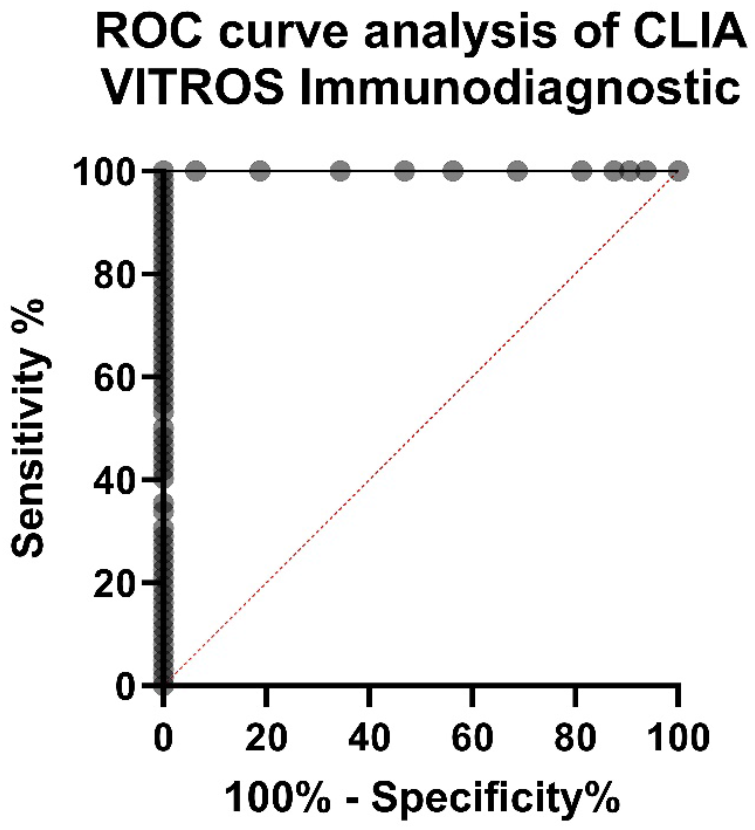
Graphical representation of ROC curve analysis of the CLIA assay by VITROS Immunodiagnostic Products. The results obtained with the pre-pandemic samples and the group of COVID-19 patients were used for the analysis. Results showed an area under the curve (AUC) of 0.9854, sensitivity of 98.41% and specificity of 93.75%.

The diagnostic performance of the test was also evaluated by making comparisons of the results obtained with the same samples, using other direct (qRT-PCR) and indirect (lateral flow chromatography and ELISA) diagnostic techniques. The results of the concordance between tests obtained are shown in Table 4. With respect to ELISA, comparisons reached a positive percentage of agreement (%PPA) of 91.3 (CI_95%_ 84.0-95.5), negative percentage of agreement (% NPA) of 97.7 (CI_95%_ 96.3-98.6), overall percentage of agreement (% OPA) of 97.7 (CI_95%_ 96.3-98.6) and Kappa index of 90.4 (CI_95%_ 85.8-94.9). In contrast, lateral flow chromatography comparisons reached a %PPA of 54.9 (CI_95%_ 47.2-62.3), % NPA of 97.9 (CI_95%_ 96.4-98.8), % OPA of 88.4 (CI_95%_ 85.9-90.5) and Kappa index of 61.1 (CI_95%_ 53.8-68.4).

**Table 4.**
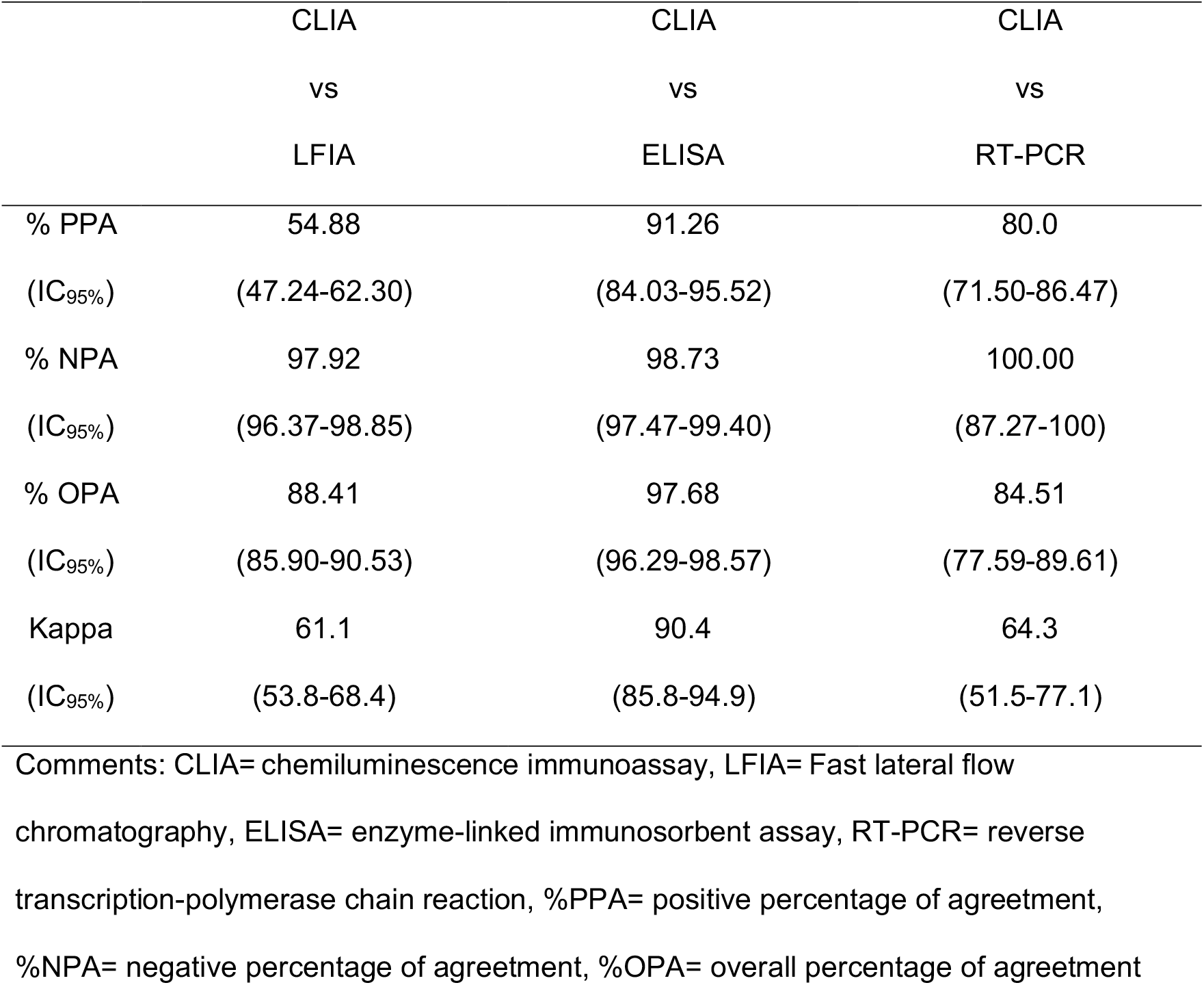
Diagnostic performance of the CLIA test VITROS Immunodiagnostic Products Anti-SARS-CoV-2.

## DISCUSSION

The significant impact of the SARS-CoV-2 emergence in public health and the massive vaccination programs carried out to limit the COVID-19 pandemic justifies extensive epidemiological studies using serological tests (11,12) to survey virus spread and assess how vaccine-induced immunity progresses in various populations and settings (13). However, due to the large volume of tests generated it is important to assess the clinical performance of the diverse immunoassays available, particularly using clinical samples from the population where the tests are implemented (14–17).

In this study, VITROS Immunodiagnostic Products Anti-SARS-CoV-2 chemiluminescence test obtained a sensitivity of 98.41% and a specificity of 93.8%. Based on the sensitivity and specificity calculated, in addition to the seroprevalence for COVID-19 reported for the study period (18%), the positive (PPV) and negative (NPV) predictive value was 77.6% and 99.6%, respectively. Sekirov et al 2021 (18) also performed a validation study of CLIA test including the one analyzed here, with in a set of samples from British Columbia, finding 100% and 98.5% of sensibility and specificity, respectively. Both studies report a high performance, however, without reaching 100% of the yield reported by manufacturer. Some authors recommended a sensitivity of 95% or more and a specificity of 99.5% or more based on samples obtained 14 days or more after symptom onset (19). Moreover, using a single cohort of samples tested on two more analytical platforms (ELISA and LFIA), the level of agreement was almost perfect with ELISA. Finally, it is important to highlight the use of serologies test (CLIA, ELISA and LFIA) for screening due to its high % NPA, useful for population studies and ruling out true negatives.

In clinical study samples (HCW, BD, and COVID-19 patients) we found a percentage of reactivity of 1.5%, 3.5% and 80.0%, respectively. The anti-SARS-CoV-2 antibody seroprevalence have been found to vary between different sites, countries, and epidemiological situations at the time of sampling. For the same period of time as our study, some studies reported high seroprevalence (20–23), while other studies (24–26), like ours, showed a low seroprevalence. Among groups, the proportion of women, ages, and reported chronic illnesses varied widely. Gruji′c et al 2022 (27) analyzed different demographic and clinical factors associated with reactivity of an anti-SARS-CoV-2 antibodies test in Serbian convalescent plasma donors, finding that variables like male sex, older age, hypertension and severe COVID-19 were linked with high anti-SARS-CoV-2 reactivity. On other hand, HCW are at a particular high risk of SARS-CoV-2 infection due to direct and indirect exposure to COVID-19 patients and aerosol-generating procedures (28,29). In line with our findings, Dusefante et al. 2022 (30) found no association between HCW and an increased occupational risk of SARS-CoV-2 infection.

## CONCLUSION

Antibodies detection tests are fundamental to epidemiological surveliiance of various infectious-related disorders including COVID-19, especially in light of the serious nature of the disease and its worldwide prevalence. In the present study, we found that the VITROS Immunodiagnostic Products Anti-SARS-CoV-2 chemiluminescence test is suitable for the detection of SARS-CoV-2 specific antibodies in a population-level screening.

## Data Availability

All data produced in the present work are contained in the manuscript

## DATA AVAILABILITY STATEMENT

All data generated and analysed in the presented study are included in this published article (and its supplementary files).

## AUTHORS’ CONTRIBUTIONS

AV conceptualized and designed the study, drafted and revised the manuscript for intellectual and scientific content; DL and AC performed CLIA test; LM contributed with the field study and clinical sample collection; GR analyzed the data, and GR and GB drafted and revised the manuscript for intellectual and scientific content. All authors reviewed the manuscript and agreed to the final manuscript version.

## ACKNOWLEDGEMENTS

We thank Dr. Amador Goodridge from INDICASAT-AIP for the access to pre-pandemic samples, also we thank Mgter. Erika Cano and Ana Julia Salerno of Ortho Clinical Diagnostics Panama for their support in the donation of the reagents from Ortho Clinical Diagnostics Panama.

## SUPLEMENTARY DATA

**Suplementary table 1.**
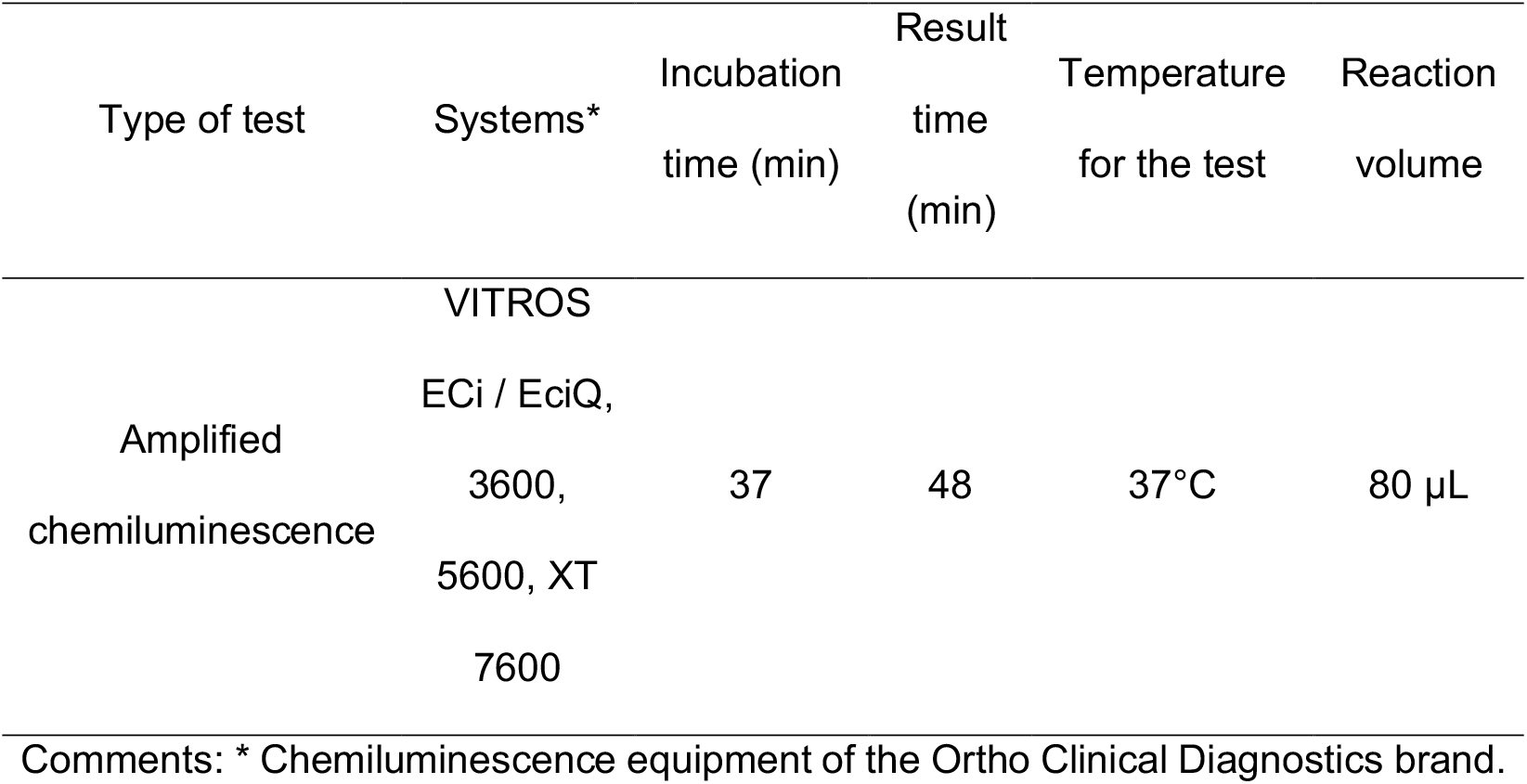
Summary of the general specifications for VITROS Immunodiagnostic Products Anti-SARS-CoV-2 Total test

**Supplementary table 2.**
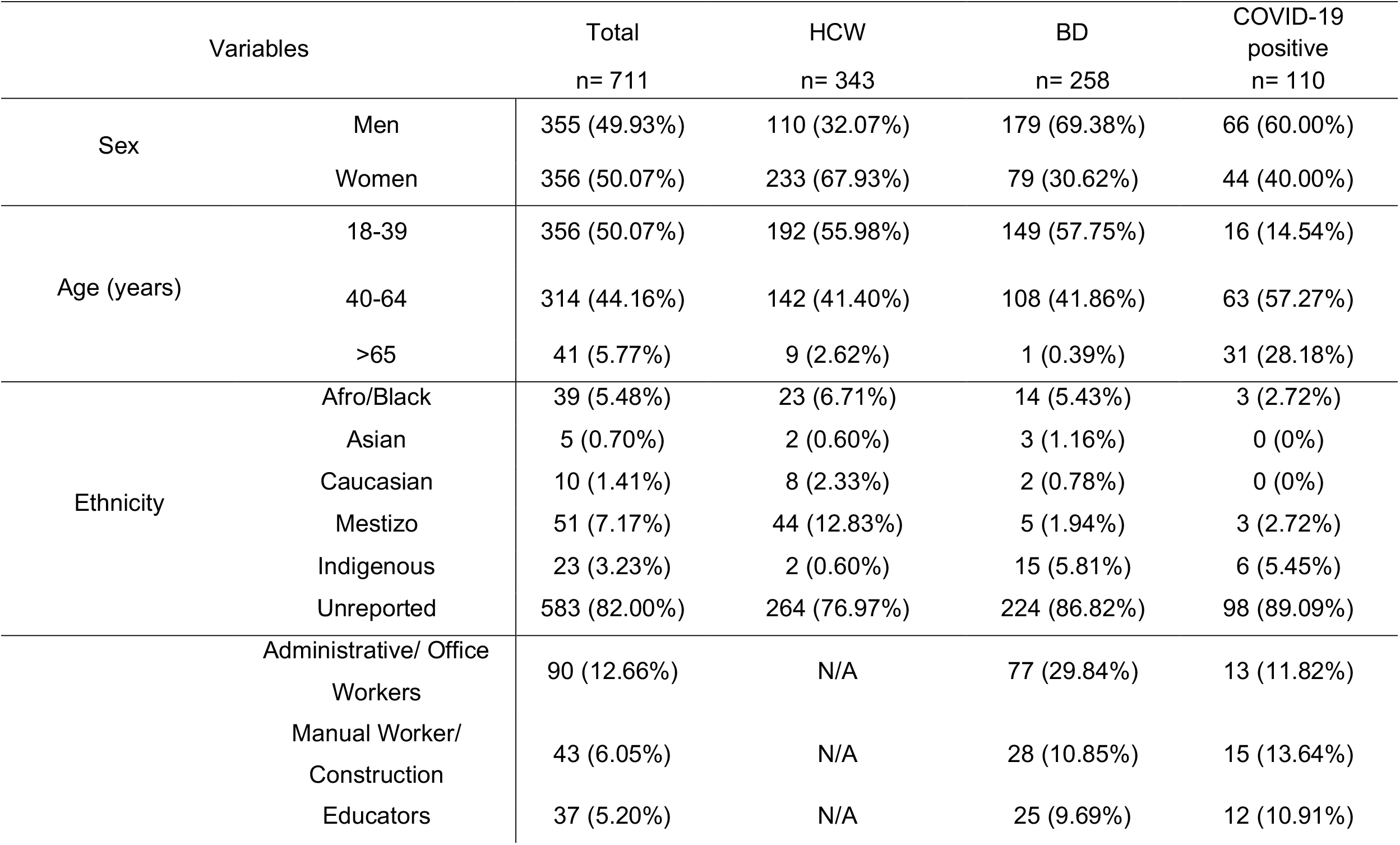
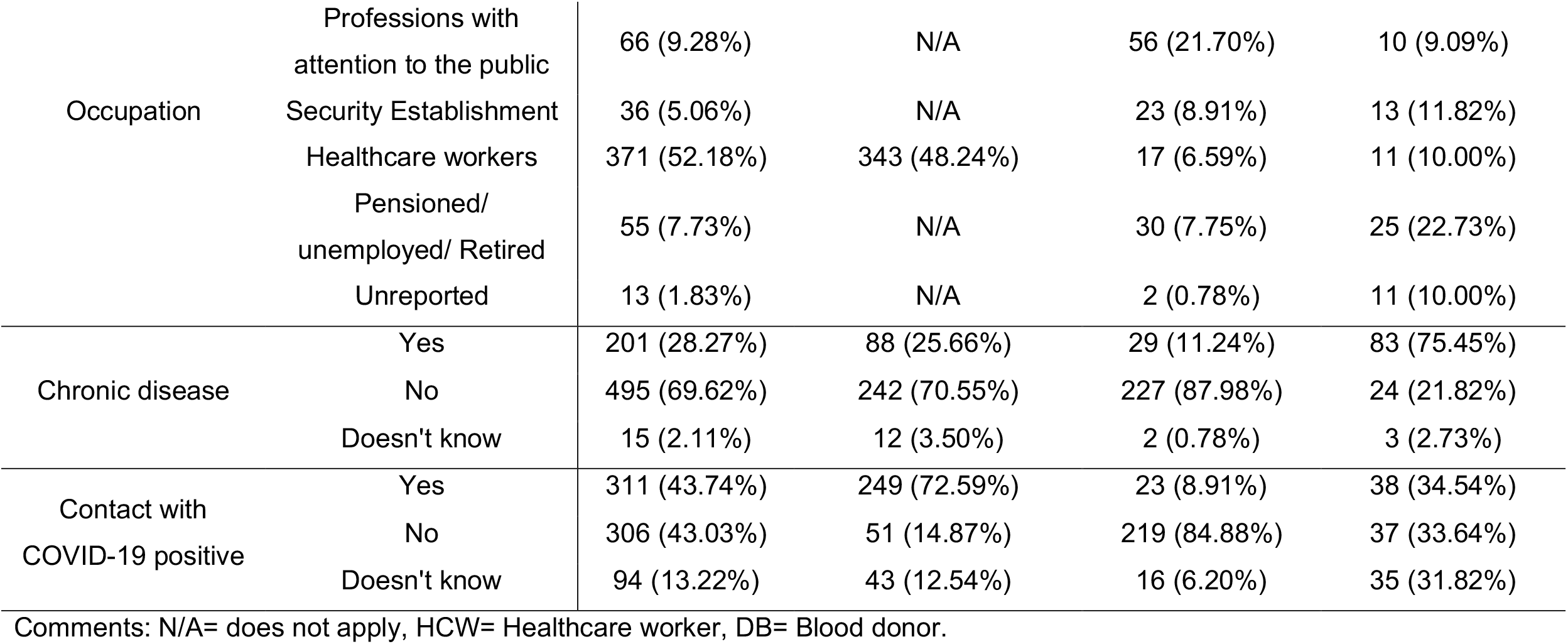
Characterization of the study population.

